# Assessing optimal time between doses in two-dose vaccination regimen in an ongoing epidemic of SARS-CoV-2

**DOI:** 10.1101/2021.07.28.21261200

**Authors:** Leonardo Souto Ferreira, Otavio Canton, Rafael Lopes Paixão da Silva, Silas Poloni, Vítor Sudbrack, Marcelo Eduardo Borges, Caroline Franco, Flávia Maria Darcie Marquitti, José Cássio de Moraes, Maria Amélia de Sousa Mascena Veras, Roberto André Kraenkel, Renato Mendes Coutinho

## Abstract

The SARS-CoV-2 pandemic is a major concern all over the world and, as vaccines became available at the end of 2020, optimal vaccination strategies were subjected to intense investigation. Considering their critical role in reducing disease burden, the increasing demand outpacing production, and that most currently approved vaccines follow a two-dose regimen, the cost-effectiveness of delaying the second dose to increment the coverage of the population receiving the first dose is often debated. Finding the best solution is complex due to the trade-off between vaccinating more people with lower level of protection and guaranteeing higher protection to a fewer number of individuals.

Here we present a novel extended age-structured SEIR mathematical model that includes a two-dose vaccination schedule with a between-doses delay modelled through delay differential equations and linear optimization of vaccination rates. Simulations for each time window and for different types of vaccines and production rates were run to find the optimal time window between doses, that is, the one that minimizes the number of deaths.

We found that the best strategy depends on an interplay between the vaccine production rate and the relative efficacy of the first dose. In the scenario of low first-dose efficacy, it is always better to apply the second dose as soon as possible, while for high first-dose efficacy, the optimal window depends on the production rate and also on second-dose efficacy provided by each type of vaccine. We also found that the rate of spread of the infection does not affect significantly the thresholds of the optimal window, but is an important factor in the absolute number of total deaths. These conclusions point to the need to carefully take into account both vaccine characteristics and roll-out speed to optimize the outcome of vaccination strategies.

## I. Introduction

As the implementations of SARS-CoV-2 vaccination programs evolve in several countries around the world, governments must decide how to allocate doses among the population during the epidemic, usually prioritizing the most at-risk groups, such as healthcare workers, people with comorbidities, and older adults. In a context of limited vaccine supply, an optimized dose allocation strategy is critical to effectively immunize the population whilst achieving the best reduction in hospitalizations and deaths. On this basis, some countries considered partially protecting a greater percentage of the population by administering the first dose more widely at the expense of delaying second doses, and thus having less fully vaccinated individuals [16, 20, 28].

Vaccination programs can have different goals depending on the context and characteristics of both the disease and the available vaccines. The immunization program against Covid-19 has focused on reducing the burden of hospitalizations and deaths, which is justified given the risk of health care system collapse and the lack of vaccine supplies to quickly reach the high coverage required to substantially reduce infections. Another strong argument for that strategy is that many of the current vaccines are highly effective against hospitalization and death, but not in preventing new infections. For instance, AZD1222 (Oxford/AstraZeneca) has shown an efficacy of 86% (95% CI: 53-96%) against hospitalization [34], but 59.9% (95% CI: 35.8-75.0%) effective against new infections measured by nucleic acid amplification-positive tests [36]; for CoronaVac (Sinovac) this contrast is even starker with 83.7% (95% CI: 58.0-93.7%) efficacy against severe cases but only 50.7% (95% CI: 35.9-62.0%) for mild symptomatic cases as outcome [24]. Even BNT162b2 (Pfizer/BioNTech), which showed high efficacy against infections in the first trials [25] may provide a lower protection against new variants, as exemplified by the variant B.1.617.2 (also known as Delta) [6]. In this context, vaccination programs in many countries have prioritised the groups most at risk of developing severe disease, namely high exposure individuals (*e*.*g*. health care professionals), people with aggravating conditions, and older adults [29, 21].

Mathematical models in epidemiology have been widely used to assess optimal vaccination strategies for a variety of communicable diseases [4, 11, 12, 13, 37]. In a previous study on influenza viruses, Matrajt et al. [19] explored how two different vaccination strategies (single dose or two dose) could be integrated into pandemic control plans of a limited vaccination supply. They conclude that the best strategy depends on the level of partial protection introduced by a single dose, but the study is limited to pre-pandemic vaccination scenario and does not capture vaccination roll-out during a pandemic.

Mass vaccination strategies have been proposed as the main approach to tackle the spread of SARS-CoV-2, complementing or replacing non-pharmaceutical interventions (NPIs). SIR-like models have pointed to the possibility of securely relaxing NPIs months after vaccination campaigns, depending on the rate of vaccination, as shown by Kraay et al. [17]. The same work also suggests that, given a low roll-out rate scenario, one-dose strategies allow safe relaxation of NPIs sooner if the first dose corresponds to more than 80% of the vaccination protection (vaccination parameters based on BNT162b2). Although one-dose vaccination allows earlier relaxation of NPIs, it requires a slower transition to pre-pandemic levels for more vaccines to be delivered. The one-dose scheme is modelled as an effectively weaker vaccine, with no second doses being considered in the long run.

In this work we aim to find the optimal time-window between doses under different scenarios of relative efficacy of the first dose and infection rate in the population. We assess these scenarios for the three most used vaccine platforms, namely inactivated virus, viral vector of adenovirus, and mRNA-based vaccines. Given that most vaccination protocols requires a second dose, we introduce a mathematical model that rigorously describes a two-dose vaccination scheme and optimal allocation of vaccines through delay differential equations. We explore the consequences of different strategies regarding the time-window between the two vaccine doses. The distribution of available vaccines for the first and second dose is optimized for each between-doses time interval, assuming a constant production rate. The partial protection offered by single-dose vaccination compared to double-dose vaccination is controlled by a single relative efficacy parameter. We find that the best strategy depends on the interplay between the vaccine production rate and the single-dose relative efficacy, with the effective reproduction number being an important factor for the reduction of deaths.

## II. Methods

To investigate the impact on total hospitalizations and mortality of delaying the second dose in a two-dose vaccination roll-out during an ongoing epidemic, we built a model that takes into account the severity of the disease, age classes (the main risk factor for severity and death), and vaccination status.

Our model consists of an **SEIR** model extended to account for asymptomatic, severe/hospitalized, and deceased individuals, thus being named **SEAIHRD**, corresponding to susceptible, pre-symptomatic, asymptomatic, mildly symptomatic, severely symptomatic/hospitalized, recovered, and deceased classes respectively. Each epidemiological class comprises three age classes, namely children and teenagers (0-19), adults (20-59), and older adults (60+) classes. To account for vaccinated individuals with one or two doses, we have duplicated this set of classes for individuals who received one dose and two doses, which have different epidemiological parameters, as discussed below. The model structure is shown in Fig. 1 and the model equations are presented in the supplementary material (SM).

**Fig. 1:**
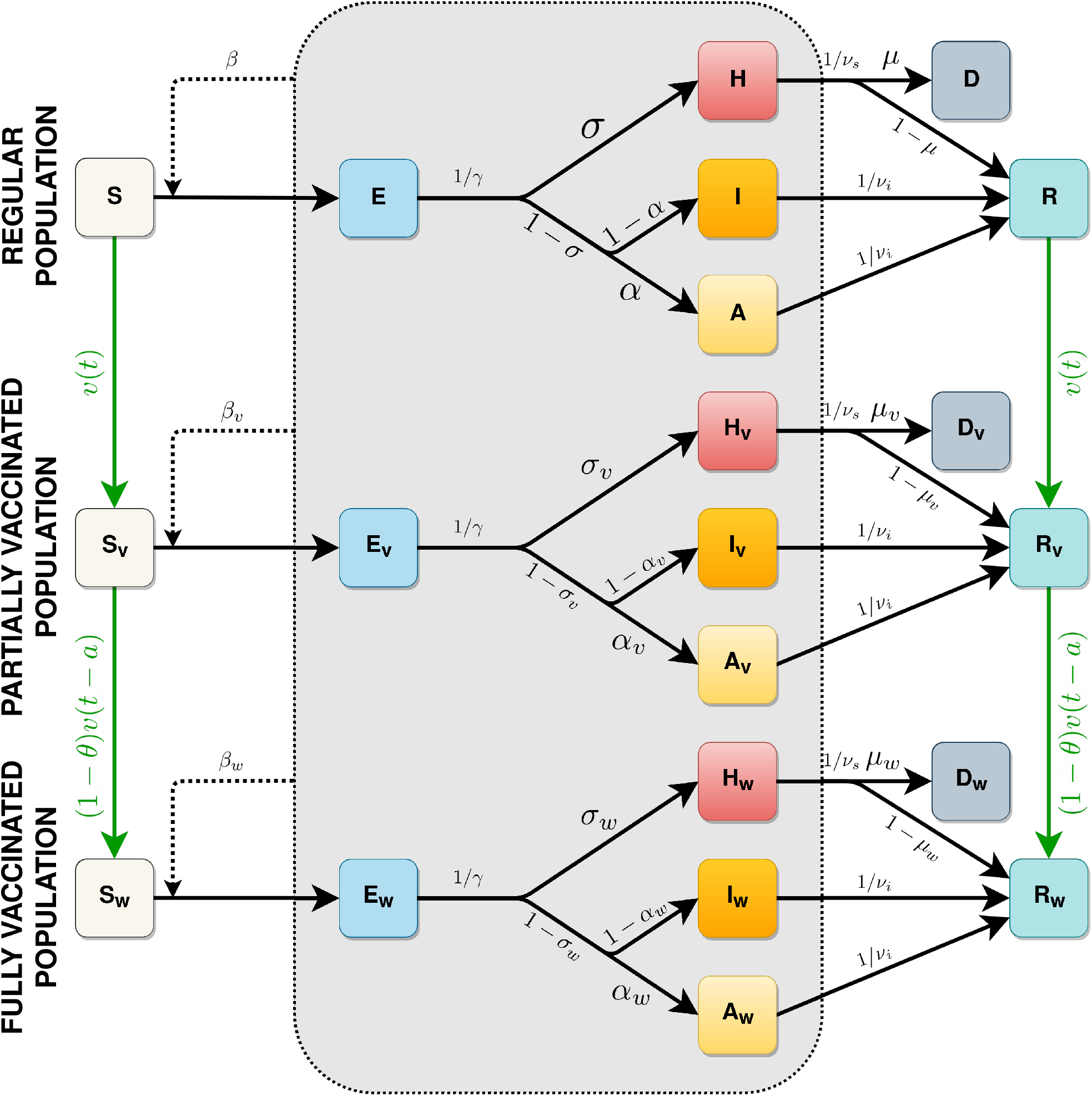
Diagram representing the full model structure. Subscripts *v* and *w* indicate the first- and second-dose vaccinated classes, respectively. Black arrows indicate transitions between epidemiological stages, green arrows indicate vaccination. All classes pictured inside the gray box are infectious. Because epidemiological progressions happen at time-scales shorter than those related to vaccine effects, infectious classes are not vaccinated in the model.

The demographic characteristics of the population were based on the State of São Paulo, Brazil. The basic epidemiological parameters come from current literature and are detailed in the SM. The daily contact rates between age classes are based on the matrices projected by Prem et al. [26], from which we use the Brazilian all-locations average for simplicity, reducing the age compartments using the same approach as in [8] (Supplementary material, section 3).

We assume a “leaky” vaccination effect, in which vaccinated individuals receives partial protection, in contrast to the “all-in” model where part of the vaccinated individuals receives full protection. Bubar et al. [7] has shown that the outcome of these models do not differ substantially and the leaky model is simpler to understand and to implement. The main parameters that describe the vaccination dynamics are the vaccine’s protective effect against (i) acquiring the disease, (ii) developing symptoms, (iii) developing severe symptoms (i.e., leading to hospitalization), and (iv) death. We present our analysis using sets of parameters that represent three types of vaccine developed for Sars-Cov-2; inactivated virus (CoronaVac, from Sinovac), adenovirus-based vaccine (AZD1222, from AstraZeneca-Oxford) and mRNA (BNT162b2, from Pfizer-BioNTech). The parameters for each vaccine are shown in Table I.

**TABLE I:**
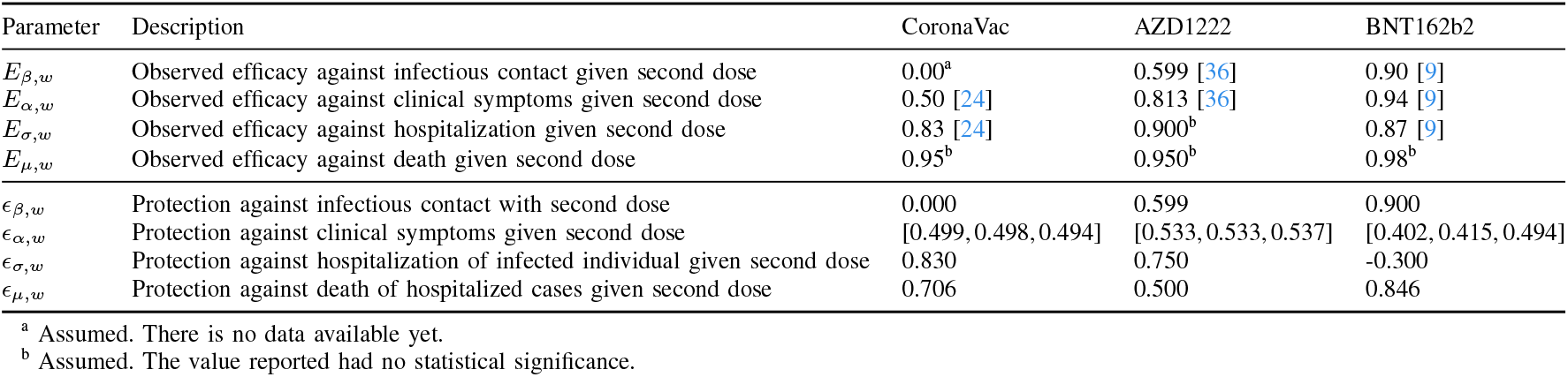
Parameters related to vaccine efficacy. The observed efficacies are obtained from trials or effectiveness studies. The parameters of protection are calculated through the method explained in the appendix A1.

| Parameter | Description | CoronaVac | AZD1222 | BNT162b2 |
| --- | --- | --- | --- | --- |
| $E_{\beta,w}$ | Observed efficacy against infectious contact given second dose | 0.00<sup>a</sup> | 0.599 [36] | 0.90 [9] |
| $E_{\alpha,w}$ | Observed efficacy against clinical symptoms given second dose | 0.50 [24] | 0.813 [36] | 0.94 [9] |
| $E_{\sigma,w}$ | Observed efficacy against hospitalization given second dose | 0.83 [24] | 0.900<sup>b</sup> | 0.87 [9] |
| $E_{\mu,w}$ | Observed efficacy against death given second dose | 0.95<sup>b</sup> | 0.950<sup>b</sup> | 0.98<sup>b</sup> |
| $\epsilon_{\beta,w}$ | Protection against infectious contact with second dose | 0.000 | 0.599 | 0.900 |
| $\epsilon_{\alpha,w}$ | Protection against clinical symptoms given second dose | [0.499, 0.498, 0.494] | [0.533, 0.533, 0.537] | [0.402, 0.415, 0.494] |
| $\epsilon_{\sigma,w}$ | Protection against hospitalization of infected individual given second dose | 0.830 | 0.750 | -0.300 |
| $\epsilon_{\mu,w}$ | Protection against death of hospitalized cases given second dose | 0.706 | 0.500 | 0.846 |

<sup>a</sup> Assumed. There is no data available yet.
<sup>b</sup> Assumed. The value reported had no statistical significance.

As the required parameters may not be available in the studies for each vaccine, specially regarding the efficacy of a single dose, we simplify the problem by assuming that the efficacy for each outcome is proportional to the efficacy after the second dose by a fixed ratio, called *relative efficacy of the first dose*. This is a key parameter influencing the model outcome: if it is too small it is better to apply the second dose sooner, but the opposite should be expected if the efficacy with one dose is as large as with two doses. Thus, we vary this parameter over all reasonable values (0-100%).

As the efficacy related to a specific outcome can be caused by a multiplicative effect of efficacy related to another outcome, (for example, lower number of deaths caused by a lower number of hospitalizations), we translate mathematically the observed efficacies that come from field studies to the parameters used in the mathematical model, as described in Appendix A1. The values of parameters used in the model are given in Table I. Notice that this mathematical translation makes the protection against clinical symptoms age-dependent and also makes the protection against hospitalization for BNT162b2 negative. The latter case occurs because the efficacy against hospitalization is lower than the efficacy against infectious contact, but when the parameters of protection are multiplied as in the model’s equations (SM) the value of efficacy is recovered.

Since we aim to study vaccination roll-out during an ongoing epidemic, we set the initial conditions for recovered and infectious populations using estimated values from hospitalization data. The procedure follows closely the one presented in Coutinho et al. [8], and is described in the SM. We use epidemiological and demographic data from the state of São Paulo, Brazil, although we do not expect the results to change substantially from one region to another.

We do not explicitly implement non-pharmaceutical interventions in our study, opting instead to model different scenarios by fixing the initial value of the effective reproduction number *R*_*t*_. Thus, we computed *R*_*t*_ using the method of Next Generation Matrix (for details, see Appendix B) and chose the value of probability of infection per contact (*β*) that yields the desired value of *R*_*t*_ at the beginning of the simulation. We vary this initial *R*_*t*_ between 0.9 and 1.4, accounting for potentially more transmissible variants [8, 10] or increased contact rates due to less rigid non-pharmaceutical interventions.

The distribution of vaccines follows the priority of age groups: first older adults, then adults, and at last children and teenagers [35]. We assume that serological tests are not performed before vaccination, therefore vaccination does not depend on the individual’s previous serological status. Thus, susceptible and recovered individuals are vaccinated proportionally to the compartment’s population size. Severe and symptomatic individuals do not receive doses while infectious due to noticeable infection. We also exclude pre-symptomatic and asymptomatic from vaccination, since it would lead to unrealistic immediate benefits and there is a window between the date of application and the start of protective effects. Recovered individuals are assumed to be completely immune and, since we fix our simulation interval at 300 days, we consider that the effects of waning immunity can be negligible. Likewise, we ignore aging dynamics, since it would be relevant only at much longer time scales.

Our model specifies that the time between the applications of the first and second doses (*a*) is fixed for all individuals, which leads to a delay differential equation formulation. In contrast, constant rate models, where individuals receive the second dose at a fixed rate with an exponential distribution of times, would imply that some of them immediately receive the second dose and lead to incorrect average times between doses when vaccination rates fluctuate over time. We also allow for a proportion *θ* of individuals that take the first but not the second dose, representing abandonment; this value is fixed at 0.1 throughout our simulations (In the case of CoronaVac vaccine phase 3 trial, the rate of abandonment was 0.16 [2]).

To optimally allocate the vaccination, keeping the number of stocked vaccines as low as possible while ensuring that second doses are available, we solve a delayed optimization problem. We assume an initial stock of vaccines *V*_0_ and a constant production (or deployment) rate *p* and solve for the control variable that is the vaccination rate *v*(*t*), with an upper limit of *v*_*max*_. The full mathematical description of the problem and its solution are described in Appendix C.

To assess which conditions is best to employ a longer (up to 12 weeks) or shorter (at least 3 weeks) interval between first and second doses, we simulate the model and compute the total number of deaths across all ages and vaccination status at the end of a period of 300 days, choosing as the optimal window the one that has the lowest number of deaths. We performed this comparison by varying the relative efficacy of the first dose and also the parameters related to the current rate of infection (*R*_*t*_) and vaccine production rate.

## III. Results

The optimal vaccine roll-out with time between doses of 3 and 12 weeks is shown in Fig. 2. We can see that for reasonable parameters of production and vaccine application rate for São Paulo State, the vaccine stock can be kept close to zero (i.e. all doses can be used immediately) after about 30 days after the beginning of dose application when using a time interval of 3 weeks. For the time interval of 12 weeks, the stock is kept close to zero during the first stage of the first dose vaccination, but doses need to be stored for the application of second-doses for a small time window. In both cases, we see a pattern of alternating stages of first and second dose vaccination, while optimally keeping the stock of vaccines close to zero.

**Fig. 2:**
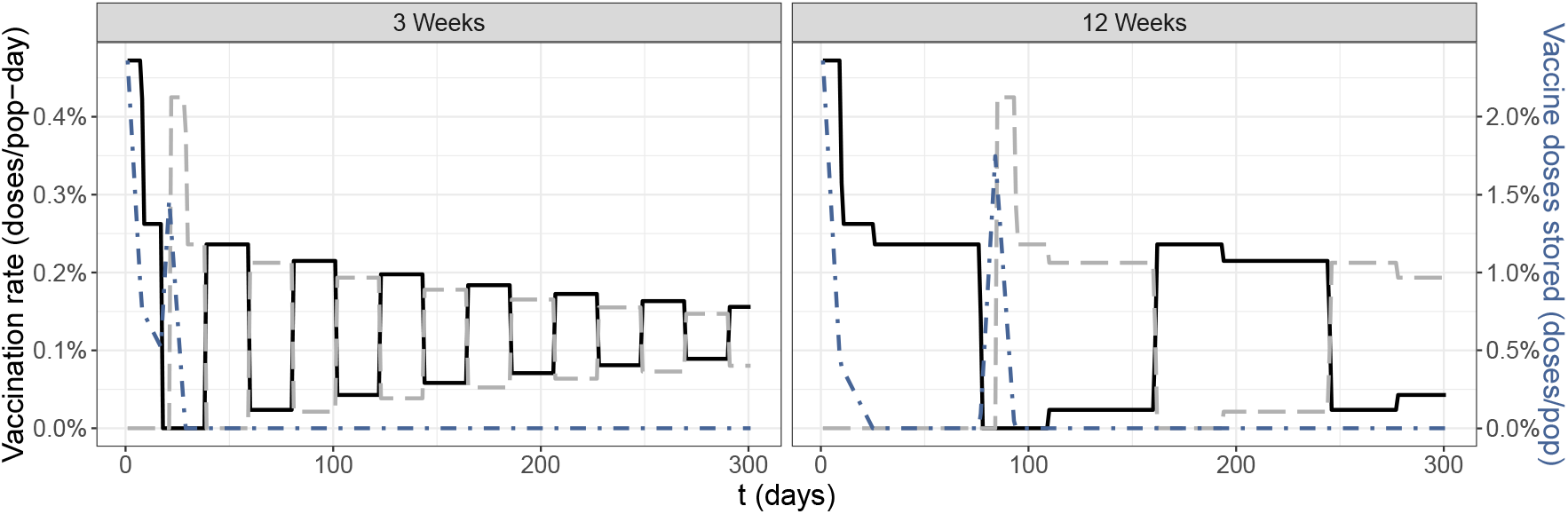
Vaccination rate as a function of time for first (solid, black) and second doses (dashed, grey) – scale is given by the left y-axis. Number of stored vaccine doses as function of time (dot-dashed, blue), scale in the right y-axis. Taking into account 3 or 12 weeks (panels) as separation between doses. *V*_0_ = 2.83% of population in doses, *ρ*(*t*) = 0.23% of population in doses/day, *v*_*max*_ = 0.45% of population in doses/day.

By varying the initial values of *R*_*t*_ for a fixed production rate (Figure 3), we see that longer periods between the first and second doses led to a lower number of deaths when the relative efficacy of the first dose alone is higher than approximately 60%, whereas smaller time intervals are better if the relative efficacy is lower than approximately 50%, and intermediary periods are the optimal interval between doses if the relative efficacy is between 50% and 60%. It is also important to note that the value of initial *R*_*t*_ does not substantially affect the value of relative efficacy where transitions between optimal windows occur, but is an important factor in the absolute value of deaths.

**Fig. 3:**
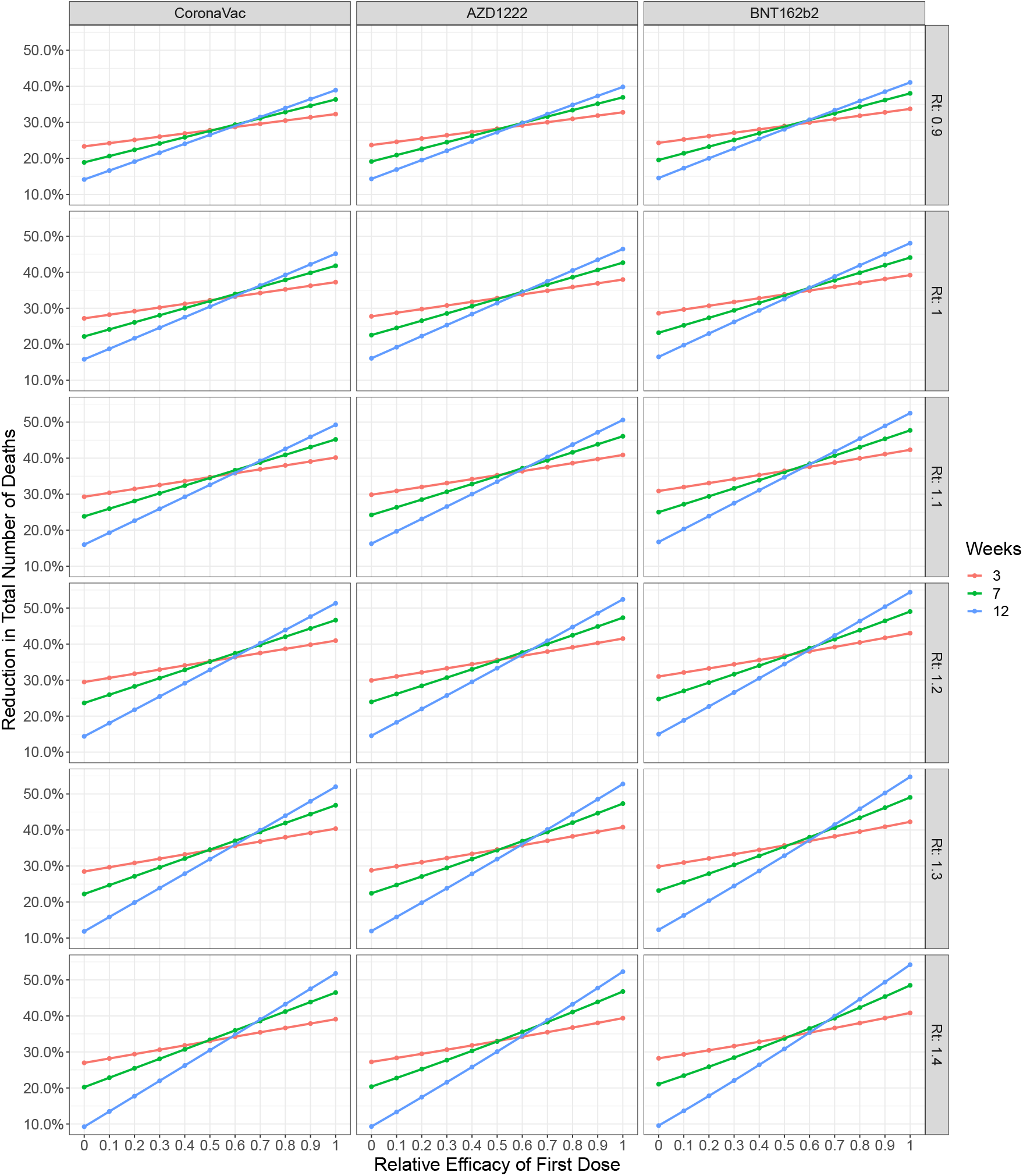
Reduction in total number of deaths as function of the first dose relative efficacy, considering three time windows between doses: 3,7 and 12 weeks (colors); varying vaccine type (columns); and effective reproduction number at the start of simulation (rows). *V*_0_ = 2.83% of population in doses, *ρ*(*t*) = 0.23% of population in doses/day, *v*_*max*_ = 0.45% of population in doses/day.

Since our results show that the optimal time window is not strongly dependant on the value of *R*_*t*_, we fixed this value at 1.1 and varied the relative efficacy and production rate. Then, we compare the optimal window relative to the reduction in deaths for several time-window intervals (Figure 4). The result is computed by simulating a coarse grid of values and then linearly interpolating a finer mesh. For all vaccines, relative efficacy of the first dose below approximately 45% indicates that regardless of the vaccine production rate, the best strategy to reduce mortality is to complete the two-dose scheme 3 weeks after the initial dose. If the production rate is low (approximately below 0.2% of doses/population-day), increasing the relative efficacy above 45% rapidly shifts the optimal time-window to increasingly larger periods, and converges to the maximum interval of 12 weeks. However, on this same range for the relative efficacy of the first dose, increasing the production rates results in a non-linear transition to shorter time-windows.

**Fig. 4:**
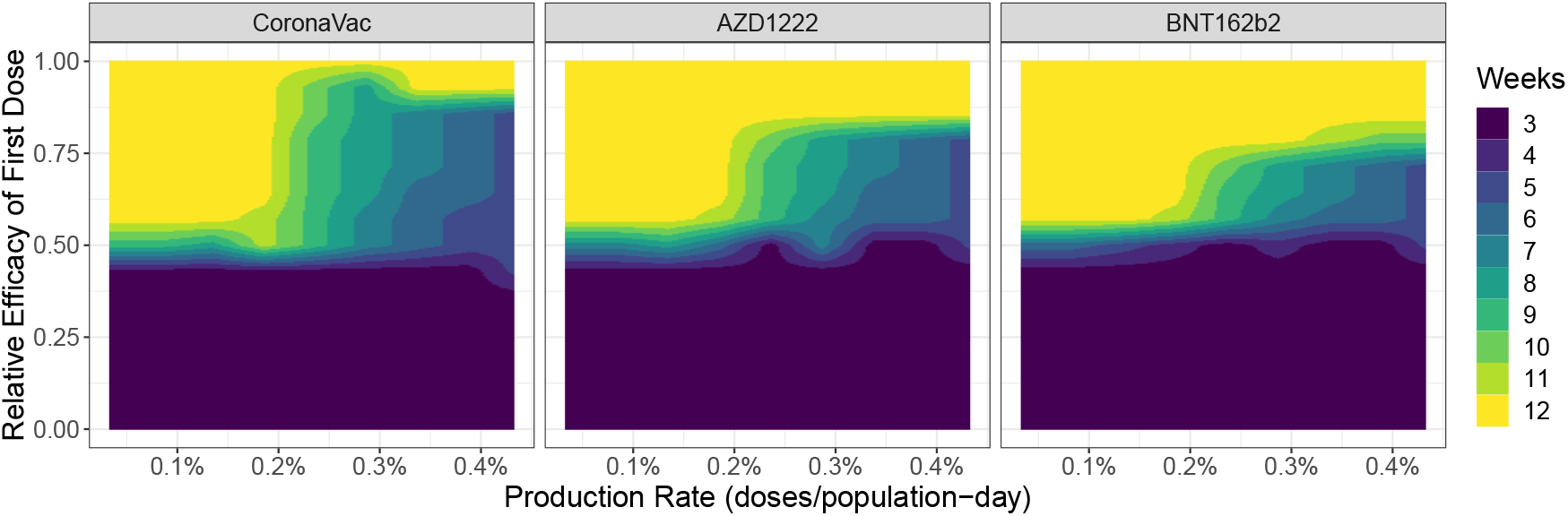
Best window (color) for reduction of deaths as function of production rate (*x* axis) and relative efficacy of first dose (*y* axis). Each panel represents a different vaccine with the respective second dose parameters. The initial *R*_*t*_ is 1.1, *V*_0_ is set to zero, vaccination rate limited to approximately 0.45% of population in doses per day.

Although it is expected that the optimal time window should always increase with higher relative efficacy, and decrease with higher production rate, the latter hypothesis is sometimes violated in the simulations (Fig. 4). Despite that, the general pattern is largely unaltered.

## IV. Discussion

Our model evaluates the optimal time windows between first and second doses for individuals in a context of limited vaccine supply in the ongoing COVID-19 pandemic. We found that the time window that best reduces the mortality depends on the interplay between two key parameters: vaccine production rate and the relative efficacy of the first dose. Within the simulated regimes with low single-dose efficacy, the resulting best strategy consistently relied on allocating the available doses to complete the two-dose scheme at the shortest time possible to provide the vaccine’s maximum protection and minimize the mortality of the population. However, when the first dose presents a higher level of relative efficacy and vaccine supply is restricted, the best strategy relied on prioritizing the allocation of doses to increase the proportion of the population receiving the first dose, resulting in a longer interval between doses to effectively allocate the use of the limited vaccine supply. Further, this interval is shortened by increasing the vaccine supply, as it progressively alleviates the conflict on dose allocation. Given that the effective reproduction number has little effect when considering the optimal window between doses, this further ensures the applicability of our results to other locations.

Another important parameter that varies across vaccines is the protection against infection, *i*.*e*. reduction in susceptibility, after the second dose, with values of 0, 0.6 and 0.9 for CoronaVac, AZD1222 and BNT162b2 respectively (See table I). This difference allows for larger delays with lower relative efficacy of the first dose, even with similar protection against deaths (0.95, 0.95, 0.98, in the same order) simply by avoiding new infections.

Given the key role of the efficacy of the first dose in deciding the best strategy to reduce mortality, it is worth considering the best available knowledge for the vaccines evaluated in this study. Preliminary results of the effectiveness of the inactivated virus vaccine CoronaVac in Chile have shown that the first-dose efficacy of that vaccine is substantially lower before the application of the second dose, with effectiveness against symptomatic infection being 17.2% (95% CI: 15.8–18.6%) [14]. This result, together with previous studies concerning inactivated virus vaccines showing that the protection against infectious contact is likely to be low [1, 32], suggests that the safest strategy concerning second dose application of CoronaVac is to apply it in three weeks, the shortest recommended interval, to achieve the largest reduction in the number of deaths. In the case of the adenovirus-based AZD1222 vaccine, Voysey et al. [36] estimated 76.7% (95% CI: 47.0-89.8%) for the efficacy against symptoms 21 days after the first dose. Albeit this estimate provides reasonably high values, the small number of events results in great uncertainty on this estimate, reflected in its wide confidence intervals. Pritchard et al. [27] corroborates those results with effectiveness against having a positive RT-PCR test of 64% after 21 days (95% CI: 59–68%) in a study in the UK, but without differentiating between AZD1222 and BNT162b2. Using this value as a proxy of first dose efficacy compared to the efficacy against symptoms post second-dose, the relative efficacy would be around 80%, which, according to our results, would suggest that delaying the second dose can be an effective strategy, especially in lower production rates. The efficacy post second-dose using a window less than 6 weeks is substantially lower (55.1%, 95% CI: 33.0-69.9%) when compared to 12 weeks (81.3%, 95% CI: 60.3-91.2%) [36], providing another argument in support of delaying the second dose of AZD1222.

For the mRNA vaccine BNT162b2, Pritchard et al. [27] have shown that its effectiveness is very high 21 days after the first dose (78%, 95% CI: 72-83%), results corroborated by a study conducted by the Joint Committee on Vaccination and Immunisation [16], from the UK. With these values, together with very high post second-dose efficacy against infectious contacts, our model results entail that postponing the application of the second dose is the best strategy. However, as this is the first known vaccine using the mRNA platform, it would be advisable to postpone the second dose only after studies assuring the efficacy of such vaccine in these conditions, as pointed out by Robertson, Sewell, and Stewart [30].

Previous studies that have evaluated the best strategies for vaccination for COVID-19 have focused on the interplay between vaccination and non-pharmaceutical interventions [7, 15, 23]. More recently, some agent-based models estimated the reduction in deaths and infections if postponing the application of the second dose: Moghadas et al. [22] found that delaying the second dose for BNT162b2 and mRNA-1273 (from Moderna) is usually the best strategy until it reaches peak values for longer intervals; Romero-Brufau et al. [31] found that the results are even better in the scenarios of low vaccine production rates. We confirm these initial results and extend the analysis to more combinations of first dose efficacy and production rates, and also consider more types of vaccine. Our model also accounts for optimal allocation of vaccination rates of first and second doses, an issue not addressed in those works.

Using a differential equations formulation, Mak, Dai, and Tang [18] compared the strategies of “hold back” (storing half of the vaccines to guarantee the second dose) and “release” (applying as much as first doses possible, while minimizing the backlog of second dose vaccination) and found that first dose efficacy is essential when deciding optimal vaccine protection coverage if delaying the second dose. Their results agree with ours but it has the limitation of not guaranteeing that the mean period with single-dose protection is preserved in a varying vaccination rate, which is only possible using a more complex formulation, such as delay differential equations as used here. They also argue that, while the “release” strategy enables better allocation of vaccines, in a real-world situation the implementation of such strategy is too complicated to manage. While we agree that this strategy adds a complication factor, we also believe that stock optimization models such as the one developed in this study could be linked to vaccination databases and, as the vaccination roll-outs to tackle the SARS-CoV-2 pandemic were centralized by the national health systems, thus allowing more efficient allocation of vaccines.

Throughout this study, we assumed that post second-dose efficacies do not depend on the time-window between doses, although evidence suggests that this is not the case for AZD1222 vaccine [36]. This simplifying assumption probably leads to a slight underestimation of the benefits of increasing the lag between doses for this vaccine. We have also assumed that first dose efficacy is constant over time, when in reality it builds up over approximately 2 weeks and wanes later on [27, 33]. This can be addressed using agent-based models [22], or age-of-infection models in a differential equations formulation, but that would greatly increase the complexity of the algorithm. This difficulty is also present when trying to compare model parameters to first dose efficacy in observational studies, as only the period after 14 days is usually considered, leaving a very short period in which events can occur. The impact of this on our conclusions can go in either direction: for shorter time windows, the efficacy in the model should be lower than the observed one, since for most of the time – the first 14 days – protection is virtually non-existent; while for longer time windows the same happens, that is, model parameters should be below measured efficacies due to waning effects of the vaccine. We currently do not have enough details of the profile of the immunological response over time to settle the issue definitively, yet we expect the pattern of the results to hold, given that we cover every possibility of relative efficacy and thus this can be compared to averaged values in time of the efficacy of the first dose.

In summary, we developed a novel approach to assess the optimal conditions to delay the application of the second dose of three different vaccine platforms for COVID-19. Our approach consisted of using a SEIR-like model coupled with a delay differential equation vaccination model and optimization of vaccine stock. We found that the first dose efficacy relative to post second dose efficacy is an essential parameter when defining the optimal window between doses, but the vaccine production rate in each country can be also decisive. Finally, we also found that those results have little dependence on the current epidemic situation at the start of the simulation. Further research in this theme would include non-constant production rate in the optimization scheme, together with starting such model in an ongoing vaccination roll-out, as well as considering lower efficacy in different age bins and other dose prioritization schemes.

## Supporting information

Equations of the model, epidemiologic parameters.

## Data Availability

The algorithm developed in this work is available in our GitHub Repository.

https://github.com/covid19br/VaxModel-paper

## Acknowledgements

We are grateful for the collaborative work of the entire group of the Observatório COVID-19 BR. In particular, we thank Monica de Bolle, Verônica Coelho and Márcia Castro for critical inputs. This work was supported by the Coordenação de Aperfeiçoamento de Pessoal de Nível Superior, Brazil (Finance Code 001 to LSF and FMDM), Conselho Nacional de Desenvolvimento Científico e Tecnológico, Brazil (grant number: 315854/2020-0 to MEB, 141698/2018-7 to RLPS, 312559/2020-8 to MASMV and 311832/2017-2 to RAK), Fundação de Amparo à Pesquisa do Estado de São Paulo, Brazil (grant number: 2019/26310-2 and 2017/26770-8 to CF, 2018/26512-1 to OC, 2018/24037-4 to SPL and contract number: 2016/01343-7 to RAK) and Swiss National Science Foundation (grant PCEFP3 181243 to VS).

## Declaration of Competing Interests

The authors declare that they have no competing interests.

## Data Availability

The algorithms used to produce the results presented here are available at https://github.com/covid19br/VaxModel-paper.

## Appendix

### A. Parameterization of the model

The parameters of the model are as follows: *β* is the probability of infection given infectious contact. *α* is the proportion of asymptomatic individuals. *σ* is the proportion of severe/hospitalized cases amongst infected individuals. *µ* is the proportion of hospitalized individuals that die. Such parameters are age-stratified unless otherwise stated and their values are given in the supplementary material.

Parameters related to vaccine efficacy are given in table I.

#### 1) Efficacy parameters computation from observed efficacies

The vaccinated classes parameters are combined with vaccine efficacies as:

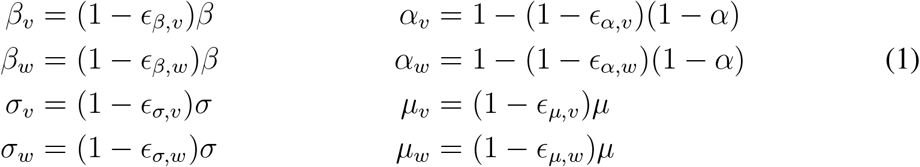

To avoid multiplicative effects in vaccine efficacies, we need to calculate the efficacy parameters from the reported values. Let us start with the risk of infection. In our model, this is given by *β*. Thus the observed efficacy against infection *E*_*β*_ is given by:

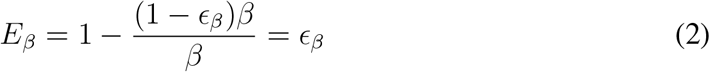

Therefore the protection against infection parameter is simply the observed efficacy. Note that we dropped the dose index as these expressions are valid for both first and second dose efficacies.

The risk of individuals being hospitalized is given by *βσ*, therefore, the observed efficacy in reducing hospitalized cases *E*_*σ*_ is then given by:

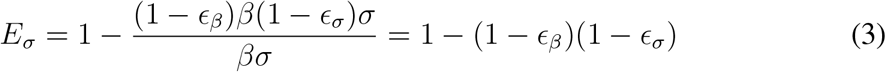

In terms of known values, the protection against hospitalization is given by:

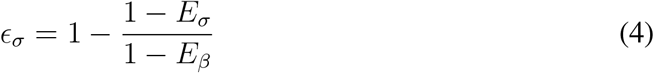

Being *µ* the proportion of hospitalized individuals that die, we have that the risk of an individual being infected and die is given by *βσµ*, therefore the observed efficacy against death *E*_*µ*_ is given by:

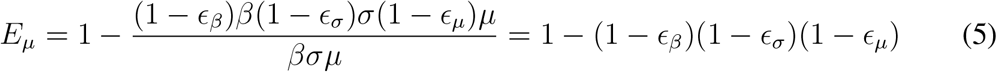

We then can obtain *ε*_*µ*_ in terms of known values:

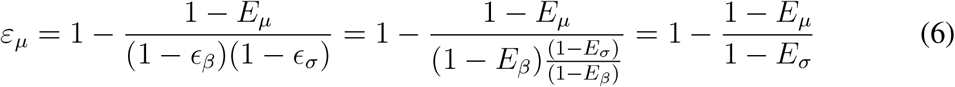

In our model, symptomatic cases are given by severe (hospitalized) and mild cases, the risk of becoming a symptomatic individual is given by *β*[*σ* + (1 *− σ*)(1 *− α*)], then the observed efficacy *E*_*symp*_ is given by:

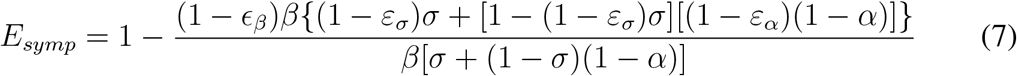

Thus

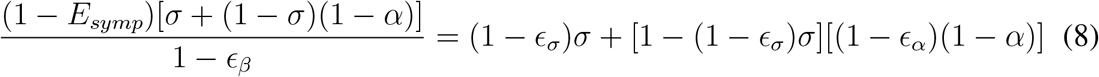

Then

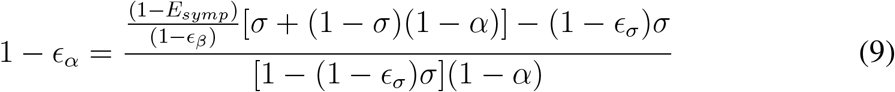

Therefore, *ε*_*α*_ is given in terms of known variables as:

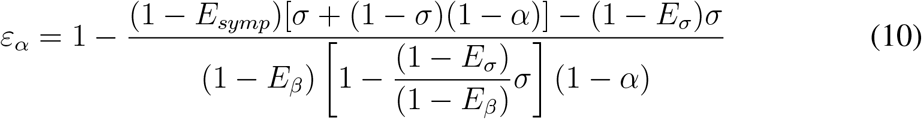

Note that 1 *− E*_*symp*_ does not multiply the whole expression.

### B. Effective reproduction number and initial conditions estimation

Both initial conditions estimation and effective reproduction number calculations go through rewriting the model in a different notation. It is a system of equations for two different groups, infected (**y**) and non-infected (**z**) populations, being **y** = (*E, A, I, H*)^*T*^, and **z** = (*S, R, D*)^*T*^. Note that none of the vaccinated classes are considered since at the initial condition no vaccine has been applied yet.

We write the system

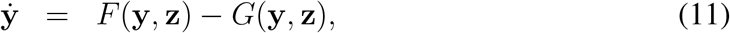

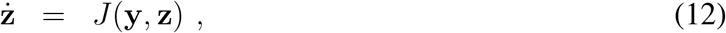

where F are all entries of new Infected, coming from classes **z**, whilst *G* accounts for the transitions within infected classes and also recovery and death from the disease. *J* accounts for the exits of the susceptible population to exposed classes, and the entrance of recovered and deceased in their respective compartments. Consider a linearization around a fixed vector 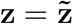, the equation for **y** becomes

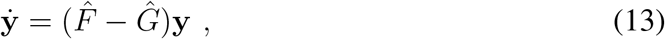

where 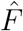 and *Ĝ* are matrices that appear from linearizing functions *F* and *G*, respectively. Remembering that each of the compartments is divided into three age sub-compartments, that is **S** = (*S*_*young*_, *S*_*adult*_, *S*_*elderly*_) and that the only entrance of new infected comes from the *β***S***λ/N* terms in the *Ė* equations, we write

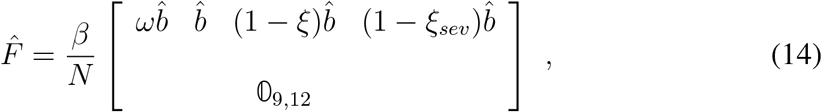

Where

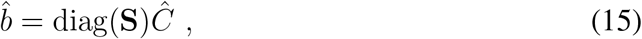

being *Ĉ* the contact matrices, from [26], *ω* the relative infectiousness of exposed individuals and *ξ* and *ξ*_*sev*_ the reductions in contacts of people that are symptomatic and hospitalized, respectively.

Now, *Ĝ* contains the terms of Exposed, *E*, developing the possible forms of disease considered in the model as the terms in its first 3 rows, while it’s main diagonal contains terms of recovery and death, writing

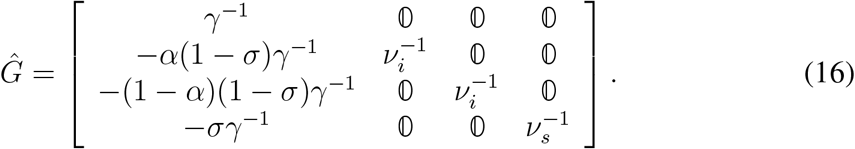

Now, *F* and *G* are important for both *R*_*t*_ calculation and initial conditions estimation. Note that for the linear problem, assuming, 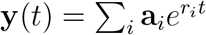, **a**_*i*_ constant vectors, yields

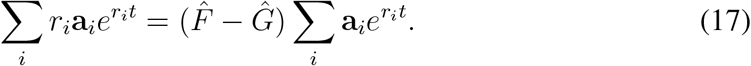

Defining *r*^*\**^ = max *r*_*i*_, **a**^*\**^ the vector associated with the exponential coefficient *r*^*\**^, and dividing the above equation by 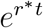, we get

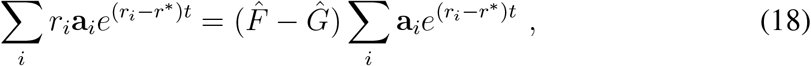

so after some time *t* elapses, the tuple (**a**^*\**^, *r*^*\**^) dominate the dynamics, and we’re left with

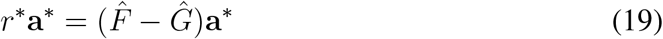

The main eigenvector of 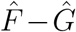, **a**^*\**^, gives a distribution of infected individuals among different classes. With hospitalizations per day data, we can fit a re-scaling factor for the eigenvector to match the term of hospital entrances (*σγ*^*−*1^*E*).

Notably, the effective reproduction calculation can be performed with 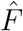 and *Ĝ* as

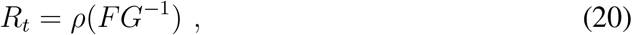

where *ρ*(*FG*^*−*1^) is the spectral radius of *FG*^*−*1^, which may be seen as the dominant eigenvalue of *FG*^*−*1^ in the simplest cases. The derivation of said result can be checked in multiple textbooks, see, for instance, chapter 6 in Allen et al. [3].

### C. Optimizing vaccination roll-out

We write a dynamical equation for the vaccine stock *V* (*t*) assuming a constant production rate *p* and a varying withdrawal rate is given by the vaccination rate, which can be chosen, that is, it is the control variable. We impose that a constant fraction *θ*^*1*^ = 1 *− θ* of the people who take the first dose will receive the second one after a period *a*, so we must be careful that the total vaccination rate is the sum of both first and second dose vaccination rates, but the control variable *v*(*t*) is the vaccination rate of first doses only. We also assume that there’s an initial stock of vaccines *V*_0_. The equation for *V* (*t*) then is:

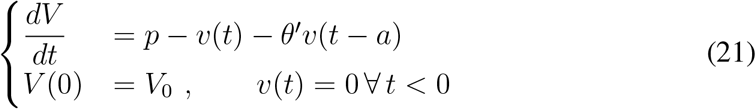

We note already that we can solve this equation, obtaining

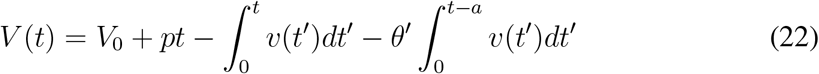

We define the optimization problem by stating the objective function to be minimized and the restrictions that the solution must obey. Since we want to use vaccine doses as quickly as possible, a reasonable goal is to minimize the stock of vaccines *V* (*t*). With that, we impose that the total vaccination rate is limited by a certain maximum value, and of course, it is positive; also, the vaccine stock *V* (*t*) is always positive. Finally, we must ensure that in the period after the simulation ends (*t > T*) there will be enough doses left to apply the second doses on those who have already taken the first dose.

These considerations lead to the following optimization problem:

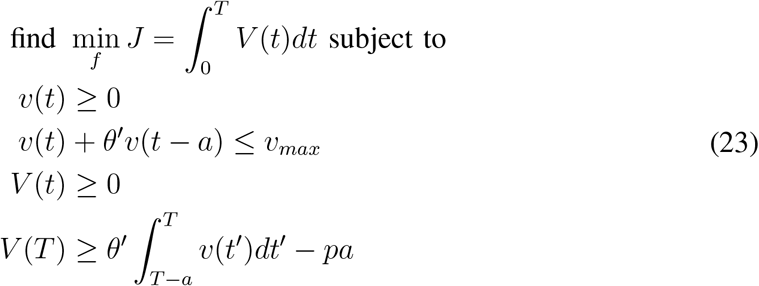

#### Solution

We can solve the problem defined by Eqs. (22, 23) using linear programming. This is feasible because the objective function and all constraints are linear functions of the control variable *f* and state variable *V* and, as Eq.(22) shows, *V* is linear on the control variable.

We first discretize the time in *n* intervals of length 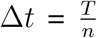, each interval ending at *t*_*i*_, *i* = 1, …, *n*, and assume that the control function will be constant over each interval (that is, a step function), with values 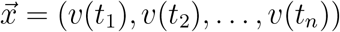. Eq.(22) then becomes

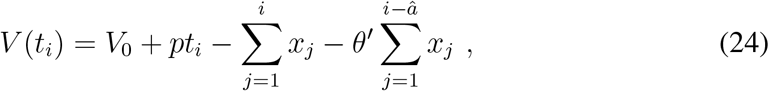

and we seek to minimize the objective function (given by Eq.(23), up to a constant) that is a linear function of 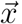, subject to the (linear) restrictions.

The discrete version of the problem becomes:

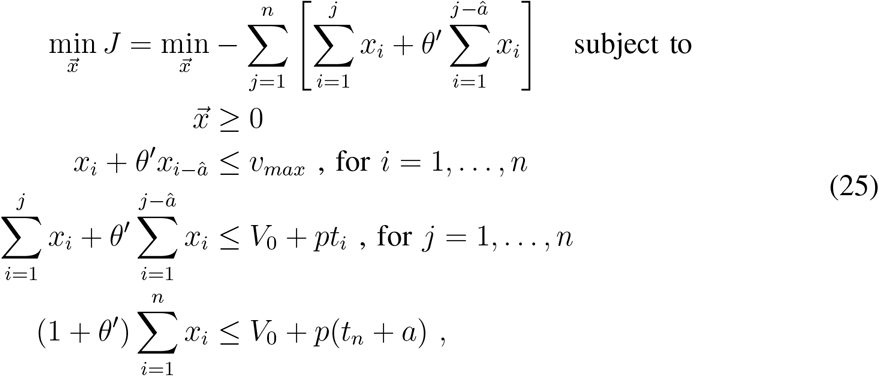

where 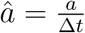 (chosen so that *â* is integer), and, to simplify notation, *x*_*i*_ is taken to be zero over values of *i* below 1.

These conditions can readily be written in matrix form and solved using standard linear programming algorithms. We implemented them in R using the package `lpSolve` [5] to solve the linear programming problem.

## Notes

### Competing Interest Statement

The authors have declared no competing interest.

### Author Declarations

Ethics approval was not necessary because this study analysed only publicly available data, not including identifiable information.

