## Supplementary material for "Assessing optimal time between doses in two-dose vaccination regimen in an ongoing epidemic of SARS-CoV-2": Equations of the model, epidemiologic parameters.

[1]*Instituto de Física Teórica, Universidade Estadual Paulista, São Paulo, Brazil*
[2]*Observatório Covid-19 BR*
[3]*Department of Ecology and Evolution, University of Lausanne, Lausanne, Switzerland*
[4]*Big Data Institute, Li Ka Shing Centre for Health Information and Discovery, Nuffield Department of Medicine, University of Oxford, Oxford, UK*
[5]*Instituto de Física 'Gleb Wataghin' and Instituto de Biologia, Universidade Estadual de Campinas, Campinas, Brazil*
[6]*Faculdade de Ciências Médicas da Santa Casa de São Paulo, São Paulo, Brazil*
[7]*Centro de Matemática, Computação e Cognição, Universidade Federal do ABC, Santo André, Brazil*

### A. *Model Equations*

To model the virus dispersal in the population, we assume that asymptomatic individuals have equal infectiousness compared to symptomatic ones, while pre-symptomatics have reduced infectiousness given by $\omega$. To model behaviour, we assume that symptomatic individuals isolate themselves at some degree, reducing their contacts by $\xi$, individuals with severe disease have greater isolation $\xi_{sev}$ due to hospitalization. The daily contacts between each age class is given by the matrix $\hat{C}$ and the force of infection $\lambda$ is given below:

$$\begin{aligned}\lambda = \hat{C}[&A + \omega E + (1-\xi)I + (1-\xi_{sev})H + \\ &A_v + \omega E_v + (1-\xi)I_v + (1-\xi_{sev})H_v + \\ &A_w + \omega E_w + (1-\xi)I_w + (1-\xi_{sev})H_w]\end{aligned} \quad (1)$$

Our model does not assume a reduction in infectiousness by vaccination given the lack of data[1].

[1]We expect that this would not change the results qualitatively.

**Unvaccinated**

$$\frac{dS}{dt} = -\beta\lambda\frac{S}{N} - v(t)\frac{S}{S+R} \tag{2a}$$

$$\frac{dE}{dt} = \beta\lambda\frac{S}{N} - \frac{E}{\gamma} \tag{2b}$$

$$\frac{dA}{dt} = \frac{\alpha(1-\sigma)E}{\gamma} - \frac{A}{\nu_i} \tag{2c}$$

$$\frac{dI}{dt} = \frac{(1-\alpha)(1-\sigma)E}{\gamma} - \frac{I}{\nu_i} \tag{2d}$$

$$\frac{dH}{dt} = \frac{\sigma E}{\gamma} - \frac{H}{\nu_s} \tag{2e}$$

$$\frac{dR}{dt} = \frac{A}{\nu_i} + \frac{I}{\nu_i} + \frac{(1-\mu)H}{\nu_s} - v(t)\frac{R}{S+R} \tag{2f}$$

$$\frac{dD}{dt} = \frac{\mu H}{\nu_s} \tag{2g}$$

**Vaccinated once**

$$\frac{dS_v}{dt} = -\beta_v\lambda\frac{S_v}{N} + v(t)\frac{S}{S+R} - (1-\theta)v(t-a)\frac{S(t-a)}{S(t-a)+R(t-a)} \tag{2h}$$

$$\frac{dE_v}{dt} = \beta_v\lambda\frac{S_v}{N} - \frac{E_v}{\gamma} \tag{2i}$$

$$\frac{dA_v}{dt} = \frac{\alpha_v(1-\sigma_v)E_v}{\gamma} - \frac{A_v}{\nu_i} \tag{2j}$$

$$\frac{dI_v}{dt} = \frac{(1-\alpha_v)(1-\sigma_v)E_v}{\gamma} - \frac{I_v}{\nu_i} \tag{2k}$$

$$\frac{dH_v}{dt} = \frac{\sigma_v E_v}{\gamma} - \frac{H_v}{\nu_s} \tag{2l}$$

$$\frac{dR_v}{dt} = \frac{A_v}{\nu_i} + \frac{I_v}{\nu_i} + \frac{(1-\mu_v)H_v}{\nu_s} + v(t)\frac{R}{S+R} - (1-\theta)v(t-a)\frac{R(t-a)}{S(t-a)+R(t-a)} \tag{2m}$$

$$\frac{dD_v}{dt} = \frac{\mu_v H_v}{\nu_s} \tag{2n}$$

**Vaccinated twice**

$$\frac{dS_w}{dt} = -\beta_w\lambda\frac{S_w}{N} + (1-\theta)v(t-a)\frac{S(t-a)}{S(t-a)+R(t-a)} \tag{2o}$$

$$\frac{dE_w}{dt} = \beta_w\lambda\frac{S_w}{N} - \frac{E_v}{\gamma} \tag{2p}$$

$$\frac{dA_w}{dt} = \frac{\alpha_w(1-\sigma_w)E_w}{\gamma} - \frac{A_w}{\nu_i} \tag{2q}$$

$$\frac{dI_w}{dt} = \frac{(1-\alpha_w)(1-\sigma_w)E_w}{\gamma} - \frac{I_w}{\nu_i} \tag{2r}$$

$$\frac{dH_w}{dt} = \frac{\sigma_w E_w}{\gamma} - \frac{H_w}{\nu_s} \tag{2s}$$

$$\frac{dR_w}{dt} = \frac{A_w}{\nu_i} + \frac{I_w}{\nu_i} + \frac{(1-\mu_w)H_w}{\nu_s} + (1-\theta)v(t-a)\frac{R(t-a)}{S(t-a)+R(t-a)} \tag{2t}$$

$$\frac{dD_w}{dt} = \frac{\mu_w H_w}{\nu_s} \tag{2u}$$

The equations were numerically solved by the R package developed by FitzJohn and Hinsley [2].

#### B. *Parameterization of the model*

The parameters that do not depend on vaccination are given in Table I.

**TABLE I:** Epidemiological parameters

| Parameter | Description | Value | Source |
|---|---|---|---|
| $\gamma$ | Average time in days between being infected and developing symptoms | 5.8 | Wei et al. [8] |
| $\nu_i$ | Average time in days between being infectious and recovering for asymptomatic and mild cases | 9.0 | Cevik et al. [1] |
| $\nu_s$ | Average time between being infectious and recovering/dying for severe cases | 8.4 | SIVEP-Gripe for São Paulo State [3] |
| $\xi$ | Reduction on the exposure of symptomatic cases (due to symptoms/quarantining) | 0.1 | Assumed |
| $\xi_{sev}$ | Reduction on the exposure of severe cases (due to hospitalization) | 0.9 | Assumed |
| $\omega$ | Relative infectiousness of pre-symptomatic individuals | 1.0 | Assumed |
| $\alpha$ | Proportion of asymptomatic cases | [0.67,0.44,0.31] | Juvenile [6] Adult and Elderly [7] |
| $\sigma$ | Proportion of infectious cases that require hospitalization | [0.001, 0.014, 0.099] | Salje et al. [5] |
| $\mu$ | In-hospital mortality ratio | [0.417,0.188,0.754] | Portella et al. [4] |
